# What factors predict ambulance pre-alerts to the emergency department? Analysis of routine data from 3 UK ambulance services

**DOI:** 10.1101/2023.12.07.23299650

**Authors:** Fiona Sampson, Richard Pilbery, Esther Herbert, Steve Goodacre, Fiona Bell, Rob Spaight, Andy Rosser, Peter Webster, Mark Millins, Andy Pountney, Joanne Coster, Jaqui Long, Rachel O’Hara, Alexis Foster, Jamie Miles, Janette Turner, Aimee Boyd

## Abstract

**Objective:** Ambulance clinicians use pre-alert calls to advise emergency departments (EDs) of the arrival of patients requiring immediate review or intervention. Consistency of pre-alert practice is important in ensuring appropriate EDs response. We used routine data to describe pre-alert practice and explore factors affecting variation in practice.

**Methods:** We undertook an observational study using a linked dataset incorporating 12 months’ ambulance patient records, ambulance clinician data and emergency call data for three UK ambulance services. We used LASSO regression to identify candidate variables for multivariate logistic regression models to predict variation in pre-alert use, analysing clinician factors (role, experience, qualification, time of pre-alert during shift), patient factors (NEWS2 score, clinical working impression, age, sex) and hospital factors (receiving ED, ED handover delay status).

**Results:** From the dataset of 1,363,274 patients conveyed to ED, 142,795 (10.5%) were pre-alerted, of whom only a third were for conditions with clear pre-alert pathways (e.g. sepsis, STEMI, major trauma). Casemix (illness acuity score, clinical diagnostic impression) was the strongest predictor of pre-alert use but male patient gender, clinician role, receiving hospital, and hospital turnaround delay at receiving hospitals were also statistically significant predictors, after adjusting for casemix. There was no evidence of higher pre-alert rates in the final hour of shift.

**Conclusions:** Pre-alert decisions are determined by factors other than illness acuity and clinical diagnostic impression. Research is required to determine whether our findings are reproducible elsewhere and why non-clinical factors (e.g. patient gender) may influence pre-alert practice.

## Introduction

### Background

Ambulance Clinicians use pre-alert (prenotification) calls to inform receiving Emergency Departments (EDs) of the arrival of a critically unwell or rapidly deteriorating patient who they believe requires urgent review and/or time-critical treatment immediately upon arrival. Pre-alerts enable the receiving ED to prepare for the patient’s arrival, including actions such as activating a trauma team, requesting specialist support, preparing specialist equipment, or ensuring availability of a resuscitation bay.^1^ The use of pre-alerts in pathways for certain conditions (e.g. stroke, ST-elevation myocardial infarction (STEMI), major trauma) is now well established but pre-alerts are also recommended for other physiological criteria or specific conditions, or patients judged to be ‘seriously injured or critically ill’.^2–4^ Pre-alerts can lead to earlier initiation of time-critical treatment, improved processes and better clinical outcomes for patients in specific patient groups including stroke, STEMI and sepsis.^5–12^

### Importance

There are concerns that a lack of effective policies for pre-alerting may lead to inconsistent response and suboptimal patient care.^3^ Overuse or inappropriate use of pre-alerts may lead to patient safety risks due to EDs diverting resources from other critically ill patients.^1,6,7,13^ Concerns about pre-alerts being used to bypass ambulance queues outside overcrowded EDs have been raised, with suggestions that pre-alert thresholds may be related to how busy ambulance clinicians believe the ED may be, which is particularly problematic in the context of overcrowded EDs.^14,15^ Similarly, there are concerns about ‘pre-alert fatigue’ where overuse of pre-alert calls leads to ED staff placing less value on the pre-alert.^16^ Consistent and appropriate use of pre-alerting is therefore key to optimising care for ED patients, particularly in the context of increasing demand and overcrowded EDs.^3,17^

Risk averseness in ambulance clinician decision making has been shown to be influenced by levels of experience, confidence and fear of blame.^18^ Given the lack of consistency in guidance at an organisation level increasing concerns about litigation and pressures to meeting ambulance handover targets, we hypothesise that pre-alert practice is affected by factors other than patient presentation and physiology.^19^ There is some evidence of disparities in pre-alert use for stroke or STEMI, with variation based on hospital, region, and patient characteristics.^20–22^ This variation is likely to be higher for conditions that constitute high numbers of pre-alerts, yet have less clear diagnostic criteria or pre-alert pathways (e.g. suspected sepsis, unspecified medical concern). Despite pre-alerts playing a key role in the transfer of care between ambulance and ED clinicians and recognised concerns that pre-alerts can place a burden on EDs understanding of appropriateness of pre-alert practice, research to understand which patients are pre-alerted and what factors may contribute to variation in practice is still limited.^22^

### Goals of this investigation

We used routine data to describe pre-alert practice in three UK NHS ambulance services to understand which patients are receiving pre-alerts and to explore potential factors affecting variation in practice. Specifically, we aimed to understand whether there were clinician, patient and/or hospital factors that influenced pre-alert practice beyond the patient’s presenting complaint.

## Methods

### Study design and setting

In the UK, ambulance services are part of the UK NHS but organisationally independent from hospitals. Ambulance clinicians are paramedics and ambulance technicians (EMTs) who usually work without medical support. The ambulance clinician phones the pre-alert through to a dedicated number in the ED and provides information in a structured way. The ED then determines the hospital response and informs the ambulance clinician where they should bring the patient.

We developed a logic model for factors affecting pre-alerts based on a rapid review of the literature and stakeholder consultation with three UK Ambulance Service Research leads and the UK Royal College of Emergency Medicine (RCEM)/Association of Ambulance Chief Executives (AACE). The model assumes that pre-alert practice may be affected by clinician factors (role, experience, sex, time of pre-alert during shift), patient factors (age, sex, NEWS2 score, clinical working impression classified into RCEM non-physiological criteria), hospital factors (catchment ED, handover delay status at time of pre-alert) and journey time.

We obtained routine, retrospective data from three adjoining ambulance services in England, covering a total population of 15.4 million people with a wide urban/rural and demographic mix. The ambulance service sites were selected pragmatically, based on their high rates of electronic Patient Report Form (ePRF) completion and accessibility with ePRF usage rate between 90% and 100%.

We analysed 12 months’ ePRF data for all 999 calls that resulted in an ambulance transporting the patient to a hospital between 1st July 2020 to 30^th^ June 2021. We collected attending ambulance clinician data (highest grade recorded on scene), dispatch system Sequence of Event log data and shift information from Global Rostering System data from each ambulance service and linked this to the ePRF data using the Computer-aided Dispatch (CAD) ID unique incident identifier. We obtained daily statistics on ambulance handover delay status at the time of pre-alert from routine SITREP data.^23^

### Analysis

We undertook univariate analysis to describe pre-alert practice, including patient characteristics and clinical information for all conveyances with and without a pre-alert to understand which patients and clinical conditions were pre-alerted. Variable selection for the multivariable logistic regression model was performed using least absolute shrinkage and selection operator (LASSO).^24^ The LASSO process begins with a full model of all potentially relevant predictors and simultaneously performs predictor selection and penalisation during model development to avoid overfitting. Potential hospital, clinician and patient predictors considered for the logistic regression model are listed in Table 1.

**Table 1.** Potential predictors for multivariable logistic regression model.

| Hospital factors |  |
| --- | --- |
| Journey time | Time arrived at hospital - Time left scene. Handover time was used where hospital arrival time was missing. |
| Handover delay status | Obtained daily for each hospital from SITREP data, proportion of ambulance handovers >30 mins.<sup>23</sup> |
| Hospital characteristics | Hospital name. Excluded where no ED, or Hyper Acute Stroke Unit or Primary Percutaneous Coronary Intervention only. |
| Ambulance clinician factors |  |
| Length of experience | Only available for sites 1 and 2. Length of clinical experience. Consecutive time in a clinical role. |
| Role | Senior clinician, paramedic, newly qualified paramedic (NQP - <2 years since qualification), non-registered clinician (e.g Emergency Medical Technician, Associate Ambulance Practitioner) and non-registered clinical support staff.<br>Simplified role only available for site 3 - paramedic / non-registered clinician |
| Qualification status | Registered on Health and Care Professions Council as paramedic. Binary yes/no<br>Sites 2 and 3 unable to provide data. |
| Sex | Male/Female/other |
| Ethnicity | ONS categories. Excluded due to poor completion. |
| Proportion of shift | Binary variable for last hour of shift. Calculated using Time left scene - Shift start time. Unable to obtain for site 3. |
| **Patient factors** |  |
| Age | Continuous variable |
| Sex | Male / Female / Transgender |
| NEWS2* | First & last NEWS2 score (clinician input NEWS2 or calculated from physiological variables). |
| RCEM non-physiological criteria. | Clinical working impression code(s).<br>Variables used to derive non-physiological criteria for pre-alert: respiratory rate, blood pressure (systolic & diastolic), pulse rate, GCS (total, eyes, verbal motor), location description (Appendix 1).<br>Interventions (airway management; defibrillation/cardioversion/pacing; trauma-related; haemorrhage control or drug administration). |

\*NEWS2 is an early warning score based on routine physiological measures that is used in UK ambulance services and hospitals as a measure of illness acuity.<sup>25</sup>

We excluded cases from the regression model where the patient was transferred to the ED from another healthcare setting (inter-facility transfers), or who were taken to the ED with a clinical working impression of ST-segment elevation myocardial infarction or hyperacute stroke, in order to mitigate for cases who had pre-alerted as part of an ED bypass (e.g. STEMI, stroke bypass as part of a pathway). We excluded patients under 16 years of age due to different physiological/ NEWS2 thresholds.

Due to high levels of correlation between NEWS2 scores and individual physiological criteria, we only included NEWS2 (i.e. not separate observations) in the final model. In order to account for presentations that may be pre-alertable but not identified by NEWS2 (e.g. acute stroke) we also included UK RCEM/AACE non-physiological pre-alert criteria^2^, using a dichotomous variable of non-physiological criteria Y/N.

We developed two clinician role categories. A simplified clinician role variable was available for all cases (paramedic, non-registered clinician). For sites 1 and 2 where more granular data was available, clinician roles were allocated 1 of 5 categories (senior clinician, paramedic, newly qualified paramedic (NQP), non-registered clinician and non-registered clinical support staff).

We undertook the following sensitivity analyses: 1) investigating the impact of selecting first NEWS2 score versus last NEWS2 score, fitting the model with the worst score, then again using the first score with an interaction term for change in score 2) using hospital as a fixed effect with a global test for the significance of hospital overall (i.e. likelihood ratio test).

Where a calculated NEWS2 score was not available in the ePR data, we calculated the NEWS2 score from the available physiological variables. Missing data was imputed with the value zero, classifying missing as normal, unless 3 or more physiological variables were missing.

### Patient and Public Involvement

Our study Patient and Public Involvement (PPI) group included people who have lived experience of pre-alerts, either through being pre-alerted themselves or through being a carer or family member of someone who has been pre-alerted. One of the PPI group was a co-applicant on the study and they attended all of the project management meetings where the research process and analysis was discussed. In addition regular PPI group meetings were held to discuss the research and the PPI group attended a 3 hour workshop where they were asked to comment on the findings.

## Results

We included 1,363,274 conveyances in the descriptive analysis after removal of 18,668 inter-facility transfers. Baseline characteristics and characteristics of pre-alerts are presented in Table 2. The dataset included 142,795 pre-alerts, with pre-alert rates by ambulance service of 8-15%. Baseline populations were similarly matched for sex but site 3 patients were older (Table 3). Overall, 75% of clinicians were paramedics (including newly qualified paramedics and senior paramedics) and pre-alerted a higher proportion of conveyances than non-paramedics (including EMTs, student paramedics and other non-registered clinical support staff) (11.0% v 9.8%).

**Table 2:** Summary table of ambulance transports to hospital stratified by pre-alert.

| Characteristic | Site 1 Pre-alert/total (%) | Site 2 Pre-alert/total (%) | Site 3 Pre-alert/total (%) | Total Pre-alert/total (%) |
| --- | --- | --- | --- | --- |
| **Overall** | 60,549/ 413,140 (14.7%) | 51,142/623,325 (8.2%) | 31,104 /326,809 (9.5%) | 142,795/1,363,274 (10.5%) |
| **Simplified clinician role** |  |  |  |  |
| Paramedic | 46,665/305,549 (15%) | 39,342/485,978 (8.1%) | 21,302/211,206 (10%) | 107,309/1,002,733 (11%) |
| Non-paramedic | 13,884/107,591 (13%) | 11,800/137,347 (8.6%) | 9,802/115,603 (8.5%) | 35,486/360,541 (9.8%) |
| **Patient sex** |  |  |  |  |
| Male | 31,550/196,484 (16%) | 26,922/298,979 (9.0%) | 16,263/157,058 (10%) | 74,735/652,521 (11%) |
| Female | 27,871/210,636 (13%) | 23,835/319,200 (7.5%) | 14,764/168,699 (8.8%) | 66,470/698,535 (9.5%) |
| Not Specified | 0/0 (%) | 385/5,146 (7.5%) | 53/691 (7.7%) | 438/5,837 (7.5%) |
| Transgender | 15/163 (9.2%) | 0/0 (%) | 22/355 (6.2%) | 37/518 (7.1%) |
| **Working Impression** |  |  |  |  |
| Sepsis | 6,705/10,138 (66%) | 11,402/20,679 (55%) | 3,372/4,004 (84%) | 21,479/34,821 (62%) |
| Unspecified medical condition | 5,668/62,620 (9.1%) | 8,673/165,959 (5.2%) | 2,470/48,208 (5.1%) | 16,811/276,787 (6.1%) |
| Acute Stroke | 5,881/8,308 (71%) | 6,417/13,812 (46%) | 2,571/5,663 (45%) | 14,869/27,783 (54%) |
| Other | 5,791/71,923 (8.1%) | 3,848/90,264 (4.3%) | 2,134/54,066 (3.9%) | 11,773/216,253 (5.4%) |
| COVID-19 | 3,525/13,942 (25%) | 4,481/25,550 (18%) | 928/4,285 (22%) | 8,934/43,777 (20%) |
| Respiratory problem | 5,839/26,628 (22%) | 676/6,479 (10%) | 2,070/15,144 (14%) | 8,585/48,251 (18%) |
| Arrhythmia | 2,504/6,740 (37%) | 1,530/10,335 (15%) | 1,970/8,069 (24%) | 6,004/25,144 (24%) |
| Lower respiratory tract infection | 926/5,059 (18%) | 2,321/19,489 (12%) | 2,532/13,287 (19%) | 5,779/37,835 (15%) |
| Cardiac problem | 1,919/21,419 (9.0%) | 701/22,097 (3.2%) | 2,687/34,571 (7.8%) | 5,307/78,087 (6.8%) |
| Trauma: other | 1,461/13,590 (11%) | 1,677/70,944 (2.4%) | 1,776/34,630 (5.1%) | 4,914/119,164 (4.1%) |
| Convulsion | 2,516/11,135 (23%) | 1,052/8,651 (12%) | 407/3,283 (12%) | 3,975/23,069 (17%) |
| STEMI | 1,728/2,059 (84%) | 1,517/3,025 (50%) | 441/1,122 (39%) | 3,686/6,206 (59%) |
| Head injury | 1,288/12,352 (10%) | 1,286/31,482 (4.1%) | 722/13,337 (5.4%) | 3,296/57,171 (5.8%) |
| Overdose | 1,762/11,565 (15%) | 284/4,807 (5.9%) | 917/9,463 (9.7%) | 2,963/25,835 (11%) |
| COPD | 1,358/4,089 (33%) | 867/4,921 (18%) | 630/3,544 (18%) | 2,855/12,554 (23%) |
| Major trauma | 1,580/2,526 (63%) | 437/3,276 (13%) | 311/958 (32%) | 2,328/6,760 (34%) |
| Neurological problem | 673/2,664 (25%) | 0/13 (0%) | 1,554/15,514 (10%) | 2,227/18,191 (12%) |
| Unspecified infection | 326/3,108 (10%) | 429/7,010 (6.1%) | 1,465/11,482 (13%) | 2,220/21,600 (10%) |
| Acute abdominal | 1,022/26,186 (3.9%) | 381/38,647 (1.0%) | 672/24,946 (2.7%) | 2,075/89,779 (2.3%) |
| Metabolic problem | 868/3,898 (22%) | 422/4,369 (9.7%) | 678/3,757 (18%) | 1,968/12,024 (16%) |
| Other | 7,209/93,191 (7.7%) | 2,741/71,516 (3.8%) | 797/17,476 (4.6%) | 10,747/182,183 (5.9%) |
| Any RCEM Prealert criteria triggered | 40,437/98,149 (41%) | 40,250/145,989 (28%) | 22,536/80,461 (28%) | 103,223/324,599 (32%) |
| Any RCEM physiological criteria triggered | 28,990/79,956 (36%) | 29,027/117,873 (25%) | 18,911/71,907 (26%) | 76,928/269,736 (29%) |
| Any RCEM non-physiological criteria triggered | 22,566/34,267 (66%) | 24,722/57,871 (43%) | 8,658/17,209 (50%) | 55,946/109,347 (51%) |
| **Triage Category** |  |  |  |  |
| 1 | 11,423/38,678 (30%) | 7,285/47,861 (15%) | 12/382 (3.1%) | 18,720/86,921 (22%) |
| 2 | 41,974/277,608 (15%) | 34,159/334,151 (10%) | 27,409/257,389 (11%) | 103,542/869,148 (12%) |
| 3 | 6,701/85,055 (7.9%) | 8,363/199,264 (4.2%) | 3,543/65,668 (5.4%) | 18,607/349,987 (5.3%) |
| 4 | 347/10,391 (3.3%) | 247/7,780 (3.2%) | 15/1,005 (1.5%) | 609/19,176 (3.2%) |
| 5 | 104/1,408 (7.4%) | 13/408 (3.2%) | 125/2,365 (5.3%) | 242/4,181 (5.8%) |
| **Hospital Department** |  |  |  |  |
| ED | 55,703/376,831 (15%) | 48,263/574,222 (8.4%) | 27,513/280,214 (9.8%) | 131,479/1,231,267 (11%) |
| Acute e.g pPCI | 3,070/4,269 (72%) | 1,952/5,028 (39%) | 361/2,676 (13%) | 5,383/11,973 (45%) |
| Unknown | 22/1,355 (1.6%) | 632/24,326 (2.6%) | 2,518/28,691 (8.8%) | 3,172/54,372 (5.8%) |
| Ward | 1,235/14,286 (8.6%) | 295/17,767 (1.7%) | 93/3,535 (2.6%) | 1,623/35,588 (4.6%) |
| Same day e.g IUC | 519/16,399 (3.2%) | 0/1,982 (0%) | 619/11,693 (5.3%) | 1,138/30,074 (3.8%) |

**Table 3.** Summary table of ambulance transports to hospital stratified by pre-alert.

| Characteristic | Site 1 |  |  | Site 2 |  |  | Site 3 |  |  |
| --- | --- | --- | --- | --- | --- | --- | --- | --- | --- |
|  | no prealert, N = 352,591<sup>1</sup> | prealert, N = 60,549<sup>1</sup> | Overall, N = 413,140<sup>1</sup> | no prealert, N = 572,183<sup>1</sup> | prealert, N = 51,142<sup>1</sup> | Overall, N = 623,325<sup>1</sup> | no prealert, N = 295,705<sup>1</sup> | prealert, N = 31,104<sup>1</sup> | Overall, N = 326,809<sup>1</sup> |
| **Ambulance journey time** | 17 (11, 24) | 14 (9, 21) | 17 (11, 24) | 16 (11, 23) | 12 (9, 18) | 16 (11, 23) | 18 (12, 27) | 16 (10, 23) | 18 (12, 27) |
| **Percentage of hospital turnaround times >30 mins** | 8 (4, 16) | 8 (5, 16) | 8 (4, 16) | 6 (2, 15) | 7 (2, 16) | 6 (2, 15) | 10 (4, 19) | 10 (5, 20) | 10 (4, 19) |
| **Clinician length of service (years)** | 6.9 (2.5, 14.9) | 6.4 (2.4, 14.4) | 6.8 (2.5, 14.9) | 6 (4, 15) | 5 (3, 11) | 6 (4, 14) | NA (NA, NA) | NA (NA, NA) | NA (NA, NA) |
| **Patient age (years)** | 63 (39, 80) | 67 (46, 80) | 64 (40, 80) | 59 (35, 79) | 70 (51, 82) | 61 (36, 79) | 65 (41, 81) | 71 (52, 82) | 66 (43, 81) |
| **First NEWS2 score** | 1.00 (0.00, 4.00) | 5.00 (2.00, 8.00) | 2.00 (0.00, 4.00) | 2.00 (0.00, 4.00) | 6.00 (3.00, 9.00) | 2.00 (0.00, 4.00) | 1.00 (0.00, 3.00) | 6.00 (2.00, 9.00) | 2.00 (0.00, 4.00) |
| **Last NEWS2 score** | 1.00 (0.00, 3.00) | 5.00 (2.00, 8.00) | 1.00 (0.00, 4.00) | NA (NA, NA) | NA (NA, NA) | NA (NA, NA) | 1.00 (0.00, 3.00) | 5.00 (2.00, 7.00) | 1.00 (0.00, 3.00) |

<sup>1</sup>Median (IQR)

We tabulated the ambulance clinician working impressions with the highest number of pre-alerts. Most common pre-alertable conditions were sepsis (34,821 pre-alerts), unspecified medical condition/ other (28,584 pre-alerts), acute stroke (14,869 pre-alerts) or Covid-19/respiratory problem/lower respiratory tract infection (23,298 pre-alerts). Pre-alert rates varied between sites for different conditions.

Sepsis and unspecified medical conditions accounted for just over a quarter of pre-alerts (26.9%). Stroke and STEMI (which have clear pre-alert and ED bypass pathways in the UK) accounted for a further 13 %. Major trauma constituted under 2% of pre-alerts, although this rose to 4% when incorporating trauma/head injury.

### Logistic regression

After excluding cases where the ED was bypassed, patients who were under 16 years of age, or whose age was not reported, and clinical working impressions of stroke or STEMI, the final dataset included 1,129,087 records for analysis. Due to differences in clinician data availability between the three sites, we were unable to include clinician data within a single dataset for all services. As a result, we created a multivariable logistic regression model utilising combined data from sites 1 and 2 (Table 4). Table 4 presents the data including ‘ambulance service’ as a variable, which an analysis of variance (ANOVA) test suggested was a more accurate model than when ambulance service was excluded (Appendix 2).

**Table 4.** Summary of multivariable logistic regression following LASSO variable selection for ambulance service pre-alert for sites 1 and 2.

| Term | Odds Ratio (95%CI) |
| --- | --- |
| Newly qualified Paramedic | 1.3 (1.26–1.34) |
| Paramedic | 1.11 (1.08–1.13) |
| Senior clinician | 1.09 (1.03–1.15) |
| Male clinician | 0.98 (0.96–1) |
| Proportion of hospital turnarounds exceeding 30 minutes | 1.74 (1.6–1.9) |
| Patient age | 1.01 (1.01–1.01) |
| Male patient | 1.24 (1.22–1.26) |
| Patient presentation meets RCEM non-physiological pre-alert criteria | 15.85 (15.52–16.18) |
| Site 1 ambulance service | 1.94 (1.86–2.02) |
| ED1 | 1.12 (1.04–1.2) |
| ED2 | 1.57 (1.5–1.64) |
| ED3 | 0.78 (0.71–0.85) |
| ED4 | 1.43 (1.34–1.51) |
| ED5 | 0.38 (0.18–0.7) |
| ED6 | 0.84 (0.77–0.92) |
| ED7 | 0.53 (0.47–0.59) |
| ED8 | 1.21 (1.15–1.27) |
| ED9 | 0.82 (0.75–0.89) |
| ED10 | 0.92 (0.86–0.98) |
| ED11 | 0.69 (0.64–0.74) |
| ED12 | 1.2 (1.13–1.26) |
| ED13 | 1.13 (1.05–1.21) |
| ED14 | 0.81 (0.76–0.86) |
| MTC1 | 1.3 (1.25–1.36) |
| MTC2 | 1.1 (0.98–1.23) |
| MTC3 | 1.66 (1.58–1.74) |
| ED15 | 1.14 (1.08–1.2) |
| MTC4 | 1.51 (1.43–1.58) |
| MTC5 | 0.89 (0.85–0.93) |
| Other | 0.56 (0.5–0.63) |
| ED17 | 1.8 (1.73–1.88) |
| ED18 | 0.87 (0.8–0.93) |
| ED19 | 1.18 (1.12–1.25) |
| ED20 | 0.74 (0.68–0.8) |
| MTC6 | 0.81 (0.77–0.85) |
| ED21 | 0.86 (0.81–0.91) |
| ED22 | 0.84 (0.7–1) |
| MTC7 | 3.18 (0.66–10.82) |
| MTC8 | 0.9 (0.85–0.96) |
| ED23 | 0.78 (0.73–0.84) |
| ED24 | 1.52 (1.43–1.61) |
| ED25 | 2.2 (2.11–2.3) |

ED prefix denotes emergency department, MTC denotes tertiary hospital designated as a major trauma centre.

### Hospital factors

Journey time was not selected by the LASSO as a significant feature contributing to pre-alerts and so does not appear in the logistic regression. However, there was considerable variation in odds ratios (OR) between hospital sites. Major trauma centre-designated hospitals were generally associated with an increased odds of a pre-alert, although this was not universal (e.g. MTC5, MTC6, MTC8). The variation was wider in the other EDs, with ORs ranging from 0.38 (95%CI 0.18–0.7, ED5) to 2.2 (95%CI 2.11–2.3, ED25). In addition, increasing hospital turnaround was associated with increased odds of making a pre-alert (1.74, 95%CI 1.6–1.9).

### Ambulance clinician factors

While length of clinician experience was not associated with pre-alerts, newly-qualified paramedics (NQP) (typically those who have been registered for less than 2 year) had a higher OR than other clinicians (OR 1.3 95%CI 1.26–1.34 vs 1.11, 95%CI 1.08–1.13 for paramedics). Clinician sex did not appear to have a significant impact on pre-alerts, and calls within the final hour of the shift were not selected by LASSO. Staff at site 1 were more likely to pre-alert than those at site 2 (OR 1.94, 95%CI 1.86-2.02).

### Patient factors

Although pre-alerted patients are older, this appears to be explained by case mix and age appears to have a minimal impact on pre-alert decision (OR 1.01, 95%CI 1.01–1.01). Male patients had a higher OR than female (OR 1.24, 95%CI 1.22–1.26). Interestingly, NEWS2 was not selected as a feature for inclusion in the model by LASSO, but meeting the RCEM non-physiological criteria for a pre-alert was the most statistically significant predictor of making a pre-alert (OR 15.95, 95%CI 15.52–16.18).

### Limitations

The differences in pre-alert rates between the three ambulance services are likely due partly to organisational differences but also due to differences in recording rates. At site 1, pre-alert recording was mandated during the final 6-month period and therefore likely to provide an accurate estimate of true pre-alert recording rates. However, recorded pre-alerts did not increase during this period.

We are unclear whether the missing data is missing at random. Missing data may be more likely for sicker patients where the notes are likely written up after patient handover. However, although pre-alert recordings may not be missing at random due to patient condition, this is unlikely to affect other results such as clinician role, hospital attending or patient sex. Even just using data for site 1 and considering sites 2 and 3 as sensitivity analyses, results demonstrate significant variation in pre-alert rates between ambulance clinicians.

Difference in pre-alert rates between sites may be due in part to under-recording of pre-alerts, or due to differences in local protocol. Data suggests that sepsis pre-alerts are higher at site 3 than for other sites, which reflects the local protocol requiring pre-alert for any red-flag sepsis. However, it is not known whether this reflects genuinely higher pre-alerting rates or higher recording of pre-alerts.

This study was undertaken within 3 UK ambulance services and transferability may be limited for settings outside the UK. However, given known recognised variation in practice and a lack of clear protocols within other settings the level of variation identified within this study is likely to be found elsewhere. The time period for which data was collected included the second period of COVID-19 lockdown in the UK (Jan – March 2021), which reduces the potential transferability of findings. The proportion of pre-alerts due to Covid-19 or respiratory disease were likely higher within this dataset than in other years. However, this is unlikely to affect pre-alert practice for other conditions significantly.

We adjusted for case mix using the UK AACE/RCEM criteria for pre-alert. However, coding of this field required assumptions to be made, including the use of physiological parameters for non-physiological criteria, since working impression codes were not available for certain pre-alertable presentations. This coding had to be undertaken on a service-by-service basis since both provided fields and working impression codes differed between services. It is possible that this may lead to differences in categorisation in the model.

Differences in proportions of patients with sex labelled as ‘transgender’ or ‘not reported’ differed by ambulance service, suggesting that some ambulance services did not use the category ‘transgender’ within their coding and this field may not be reliable.

The analysis was exploratory in nature and not confirmatory. The aim was to explore what variables might predict the use of pre-alerts, with the aim to guide future research. Although the size of effect was different within the combined and separate models (which may be expected as these were different datasets), the different models did all show that clinical variables are the key predictors, with hospital factors, anticipated handover delay, patient sex and clinician role all being predictors.

## Discussion

Our analysis of over 1.3 million ambulance conveyances identified differences in pre-alert practice that were not attributable to case mix. We identified that pre-alert practice was affected by a combination of hospital, clinician, and patient factors. Although many pre-alerts were for key pre-alertable conditions (sepsis, stroke, STEMI, or trauma), around two-thirds of pre-alerts were for conditions that may require a higher level of clinical judgement when deciding whether to pre-alert. Within this analysis, hospital conveyed to and hospital turnaround status were key factors in pre-alert practice suggesting that concerns about anticipated response from ED may have an influence over decision making. There was some evidence that newly qualified paramedics may pre-alert more than more experienced clinicians, although the size of effect detected was small. We found no evidence that clinicians in the final hour of their shift were more likely to pre-alert.

Despite the impact and importance of pre-alerts on patient care we have not identified other literature exploring pre-alert practice and factors affecting pre-alert rates for general populations, although several studies have reported on pre-alert practice for specific conditions where the benefit of pre-alert is more clearly defined.

Difference in practice between ambulance services suggests local protocols and priorities have an impact on pre-alert practice. Boyd et al identified important differences in ambulance service guidance in the UK, with differences in physiological thresholds for pre-alert even for conditions with established care pathways with services listing between 4 and 45 conditions as suitable for pre-alert. There was also variation from the joint ambulance and ED guidance that should be used to determine whether a pre-alert is undertaken.^2^ In the US EMS criteria for pre-alert are likely to vary and practice appears to be dictated by requirements of local EDs.^3,26^ Lin et al also identified statistically significant differences in EMS prehospital notifications for stroke between hospitals and regions and concluded that disparities in EMS prenotification use occurred by state and geographic region.

Inconsistent pre-alerting within individual hospitals has been reported elsewhere, with Sheppard et al and Brown et al both identifying under-alerting of patients with suspected stroke, including finding that pre-alerts were not consistently used in suspected stroke patients, with 27% of patients who were FAST positive not pre-alerted, and 22% of patients who met their local criteria for pre-alert not being pre-alerted.^6,7^

Other studies have identified differences in patient factors affecting pre-notification for specific conditions. Blusztein et al identified male gender as an independent predictor of pre-notification for STEMI.^27^ Lin et al found that female patients were less likely to receive EMS prenotification for stroke but also identified higher likelihood of EMS prenotification for younger patients and significant ethnic disparities in prenotification, with adjusted odds ratio of pre-alert for black patients of 0.94 (CI0.92-0.97) compared with white patients.^28^ Sheppard et al did not identify any statistically different racial or sex differences in stroke pre-alerting, which could be due to the small overall sample (n=271).^7^

It is unclear whether differences identified within our study are due to case mix that was not detected within the model, implicit bias in practice or different presentation of symptoms. We were unable to explore racial differences due to poor reporting of ethnicity within the ePRF. However, our study supports findings of previous studies that report disparities in treatment based on non-clinical patient characteristics and suggests that inequalities in care exist.^29^

Our findings also demonstrate that pre-alert decision-making is affected by clinician and contextual factors, with pre-alert decisions being affected by anticipated ambulance handover delay as well as clinical experience. Weyman et al similarly identified that clinician perceptions of personal vulnerability and organisational blame in the event of a wrong decision (e.g. waiting in a queue) are likely to influence more risk-averse decision-making.^30^

## Conclusions

Given the value of pre-alerts in improving time-critical treatment for specific conditions such as stroke, sepsis, or STEMI, it is important that pre-alerts are not over-used and are used appropriately in order to prevent alert fatigue. Alert fatigue in other areas has been shown to lead to desensitisation and delayed or missing response to alerts.^16^

Considerable levels of variation in pre-alert practice across and within different ambulance services suggests that procedures and processes for pre-alerting may lack clarity and improved pre-alert protocols may be required to reduce variation. Whilst broad definitions such as ‘critically ill’ enable clinicians to use their clinical acumen to decide whether a patient requires a pre-alert, this can also enable over-use of pre-alerts and increase risk of pre-alert fatigue. There is a need to understand the reasons behind the variation in pre-alert practice identified within this analysis.

## Supporting information

Appendix 1-RCEM criteria derivations by service

Appendix 2-Summary-of-multivariable-regression

## Data Availability

The data used for this study are subject to data sharing agreements with EMAS, WMAS and YAS which prohibit further sharing of individual level data. The datasets used are obtainable from these organisations subject to necessary authorisations and approvals.

## Notes

### Competing Interest Statement

The authors have declared no competing interest.

### Funding Statement

This research was funded by the National Institute for Health and Care Research (NIHR HS&DR 131293). The views expressed in this publication are those of the author(s) and not necessarily those of the NIHR or the UK Department of Health and Social Care.

### Author Declarations

Ethical approval for the pre-alerts project has been obtained from Newcastle & North Tyneside 2 Research Ethics Committee (Ref: 21/NE/0132)

