## Appendix 1-RCEM criteria derivations by service for "What factors predict ambulance pre-alerts to the emergency department? Analysis of routine data from 3 UK ambulance services"

| RCEM criteria | Site 1 | Site 2 | Site 3 |
| --- | --- | --- | --- |
| Cardiac arrest | Working impression of cardiac arrest | Working impression of cardiac arrest | Working impression of cardiac arrest (cardiovascular); |
| Airway compromise | Any of the following interventions recorded: Oropharyngeal airway, nasopharyngeal airway, supraglottic airway device, tracheal intubation | Any of the following interventions recorded: Oropharyngeal airway, nasopharyngeal airway, supraglottic airway device, tracheal intubation | No airway adjunct data |
| Major trauma tool positive | Any of: Working impression of major trauma, Working impression of "Stabbed/shot/weapon wound", "Head injury", "Multiple injuries", "Haemorrhage/lacerations AND any of the following interventions: Tranexamic acid administration, Celox bandage, tourniquet, ketamine administration, pelvic splinting, pressure dressing,  Working impression of "Stabbed/shot/weapon wound", "Head injury", "Multiple injuries", "Haemorrhage/lacerations AND any RCEM physiological criteria outside of normal range or GCS < 14. | Any of: Working impression of "trauma: stabbing\|trauma: gunshot\|trauma: penetrating\|trauma: head injury\|trauma: multiple injuries\|trauma: wound lacerations" AND any of: Tranexamic acid administration, Celox bandage, tourniquet, ketamine administration,  Hospital department is major trauma centre,  Working impression of "trauma: stabbing\|trauma: gunshot\|trauma: penetrating\|trauma: head injury\|trauma: multiple injuries\|trauma: wound lacerations" AND any RCEM physiological criteria outside of normal range or GCS < 14. | Any of: Working impression of "gunshot (trauma);", "stab wound (trauma);", "penetrating trauma (trauma);", "multi-system trauma (trauma);", "explosive / blast injury (trauma);", "head injury (trauma);" AND tranexamic acid or ketamine administration,  Working impression of "gunshot (trauma);", "stab wound (trauma);", "penetrating trauma (trauma);", "multi-system trauma (trauma);", "explosive / blast injury (trauma);", "head injury (trauma);" AND any RCEM physiological criteria outside of normal range or GCS < 14. |
| ST-elevation MI | Working impression of ‘Cardiac STEMI’ | Hospital department is ‘ppci’ or working impression of ‘medical: stemi’ | Working impression of ‘ecg confirmed st segment elevated mi (cardiovascular);’ |
| Complete heart block or broad complex tachycardia with adverse features (shock, syncope, heart failure, myocardial ischaemia) | (Pulse rate <= 30 or >= 150 ) AND systolic blood pressure < 90. NB Children excluded | Pulse rate <=30 or >=150 and systolic blood pressure < 90 | (Pulse rate <= 30 or >= 150 ) AND systolic blood pressure < 90. NB Children excluded |
| FAST-positive stroke within timeframe for thrombolysis | Working impression of ‘Stroke – FAST positive’ | Working impression of ‘medical: stroke’ and hospital department = ‘fast +’ | Working impression of ‘stroke (neurological);’ |
| Sepsis with red flags triggering the Sepsis Trust prehospital bundle | Working impression of ‘Sepsis’ | Working impression of ‘medical: sepsis’ | Working impression of ‘sepsis (medical);’ |
| Uncontrolled seizure (still fitting) | Not possible to determine | Not possible to determine | Not possible to determine |
| Obstetric emergency e.g. maternal convulsions, shoulder dystocia or abnormal presentation of the baby | Any of: Working impression of ‘Obstetric emergency (other)’, or Working impression of ‘Obstetric – miscarriage’ and systolic blood pressure <90 | Any of: Working impression of "medical: miscarriage, medical: ectopic, medical: labour, medical: pregnancy, medical: child birth" AND systolic blood pressure < 90,  Working impression of ‘medical: eclampsia’ | Any of,  Working impression of "miscarriage (obs/gynae)", "antepartum haemorrhage (obs/gynae);", "ectopic pregnancy (obs/gynae); AND systolic blood pressure <90,  Working impression of: "eclampsia (obs/gynae); postpartum haemorrhage (obs/gynae); delivery complication (obs/gynae); |
| Life threatening asthma | Working impression of ‘asthma’ and any RCEM physiological criteria outside normal range | Working impression of ‘asthma’ and any RCEM physiological criteria outside normal range | Working impression of ‘asthma’ and any RCEM physiological criteria outside normal range |
| Uncontrolled major haemorrhage | Working impression of "Haemorrhage/lacerations", "Haematemesis", "Bleeding PR", "Bleeding PV" AND (heart rate > 130 or systolic blood pressure < 90) | Working impression of "medical: pv bleed\|medical: pr bleed\|medical: heamatemisis\|medical: gi bleed\|medical: malena" AND (heart rate > 130 or systolic blood pressure < 90) | Working impression of "gi bleed (medical);", "pv bleed (obs/gynae);" AND heart rate >130 or systolic blood pressure < 90 |
| Unconscious with a GCS motor score of less than 4 | Motor component of GCS < 4 | Motor component of GCS < 4 | Motor component of GCS < 4 |
| Overdose with abnormal physiology and possible lethality, which may require immediate intervention on arrival | Working impression of ‘Drug Overdose’ and any of:  Oropharyngeal airway, nasopharyngeal airway, supraglottic airway, tracheal intubation, systolic blood pressure < 90, heart rate >130, GCS < 8, respiratory rate <8 | Working impression of ‘medical: overdose’ and any of:  Oropharyngeal airway, nasopharyngeal airway, supraglottic airway, tracheal intubation, systolic blood pressure < 90, heart rate >130, GCS < 8, respiratory rate <8 | Working impression of "intentional drug overdose (mental health);", "accidental overdose / poisoning (medical);" AND systolic blood pressure < 90, heart rate >130, GCS < 8, respiratory rate <8 |
| Unstable vascular emergencies with clinical shock (e.g. AAA, thoracic dissection) or acutely ischaemic limb | (Working impression of ‘Vascular emergency (non AAA)’ or ‘AAA’ ) AND (heart rate >130 or systolic blood pressure <90 | Working impression of ‘medical: aortic aneurysm’ AND (heart rate >130 or systolic blood pressure <90 | Working impression of ‘aaa (cardiovascular)’ and (heart rate >130 or systolic blood pressure <90 |

Note: First and last physiological observations are included in the dataset and for all of the physiological criteria specified, if either the first or last set of observations meet the criteria, then it is assumed to be a positive result.
