## Appendix 2-Summary-of-multivariable-regression for "What factors predict ambulance pre-alerts to the emergency department? Analysis of routine data from 3 UK ambulance services"

### Appendix 2: Summary of multivariable logistic regression following LASSO variable selection for ambulance service pre-alert, stratified by inclusion of service variable for sites 1 and 2.

| Term | Service term included  OR (95%CI) | Service term NOT included  OR (95%CI) |
| --- | --- | --- |
| Newly qualified Paramedic | 1.3 (1.26–1.34) | 1.26 (1.23–1.3) |
| Paramedic | 1.11 (1.08–1.13) | 1.05 (1.03–1.08) |
| Senior clinician | 1.09 (1.03–1.15) | 1.07 (1.01–1.13) |
| Male clinician | 0.98 (0.96–1) | 0.98 (0.96–1) |
| Proportion of hospital turnarounds exceeding 30 minutes | 1.74 (1.6–1.9) | 1.96 (1.79–2.13) |
| Patient age | 1.01 (1.01–1.01) | 1.01 (1.01–1.01) |
| Male patient | 1.24 (1.22–1.26) | 1.24 (1.22–1.26) |
| Patient presentation meets RCEM non-physiological pre-alert criteria | 15.85 (15.52–16.18) | 15.69 (15.37–16.03) |
| Site 1 ambulance service | 1.94 (1.86–2.02) | NA (NA) |
| ED1 | 1.12 (1.04–1.2) | 0.78 (0.74–0.84) |
| ED2 | 1.57 (1.5–1.64) | 2.09 (2–2.17) |
| ED3 | 0.78 (0.71–0.85) | 0.54 (0.5–0.59) |
| ED4 | 1.43 (1.34–1.51) | 1.91 (1.8–2.02) |
| ED5 | 0.38 (0.18–0.7) | 0.5 (0.24–0.92) |
| ED6 | 0.84 (0.77–0.92) | 0.59 (0.55–0.65) |
| ED7 | 0.53 (0.47–0.59) | 0.37 CI:0.33–0.42 |
| ED8 | 1.21 (1.15–1.27) | 1.6 (1.53–1.67) |
| ED9 | 0.82 (0.75–0.89) | 0.57 (0.53–0.62) |
| ED10 | 0.92 (0.86–0.98) | 0.63 (0.6–0.67) |
| ED11 | 0.69 (0.64–0.74) | 0.92 (0.85–0.99) |
| ED12 | 1.2 (1.13–1.26) | 0.82 (0.79–0.86) |
| ED13 | 1.13 (1.05–1.21) | 0.79 (0.74–0.85) |
| ED14 | 0.81 (0.76–0.86) | 1.09 (1.03–1.15) |
| MTC1 | 1.3 (1.25–1.36) | 1.72 (1.65–1.79) |
| MTC2 | 1.1 (0.98–1.23) | 1.45 (1.29–1.62) |
| MTC3 | 1.66 (1.58–1.74) | 2.22 (2.12–2.33) |
| ED15 | 1.14 (1.08–1.2) | 0.8 (0.76–0.84) |
| MTC4 | 1.51 (1.43–1.58) | 1.05 (1–1.09) |
| MTC5 | 0.89 (0.85–0.93) | 1.17 (1.12–1.22) |
| Other | 0.56 (0.5–0.63) | 0.56 (0.5–0.64) |
| ED17 | 1.8 (1.73–1.88) | 2.39 (2.31–2.48) |
| ED18 | 0.87 (0.8–0.93) | 0.6 (0.56–0.65) |
| ED19 | 1.18 (1.12–1.25) | 1.58 (1.5–1.66) |
| ED20 | 0.74 (0.68–0.8) | 0.51 (0.47–0.55) |
| MTC6 | 0.81 (0.77–0.85) | 0.56 (0.54–0.59) |
| ED21 | 0.86 (0.81–0.91) | 1.15 (1.08–1.22) |
| ED22 | 0.84 (0.7–1) | 1.1 (0.92–1.32) |
| MTC7 | 3.18 (0.66–10.82) | 4.32 (0.9–14.64) |
| MTC8 | 0.9 (0.85–0.96) | 0.63 (0.6–0.66) |
| ED23 | 0.78 (0.73–0.84) | 0.55 (0.51–0.59) |
| ED24 | 1.52 (1.43–1.61) | 1.06 (1–1.11) |
| ED25 | 2.2 (2.11–2.3) | 2.96 (2.84–3.09) |

#### Analysis of variance (ANOVA) between models (service term/no-service term variable)

| Model | Residual degrees of freedom | Residual deviance | Degrees of freedom | Deviance | P value |
| --- | --- | --- | --- | --- | --- |
| Service term | 759993.000 | 419930.741 | NA | NA | NA |
| No service term | 759994.000 | 420530.951 | -1.000 | -600.210 | 0.000 |
